# Application of RNA-sequencing based predictive model for endometrial WOI in patients with recurrent implantation failure: a prospective cohort study

**DOI:** 10.1101/2024.05.26.24307954

**Authors:** Aihua He, Tianli Yang, Sijia Lu, Yangyun Zou, Cheng Wan, Jing Zhao, Nenghui Liu, Donge Liu, Yumei Li, Yonggang Wang, Bin Xu, Jie Hao, Shi Xie, Jing Fu, Hui Li, Hong Wu, Qiong Zhang, Yanping Li

## Abstract

**Background:** Accurate prediction for endometrial window of implantation (WOI) would maximize the effectiveness of assisted reproductive technology. Previously, we have established a predictive model for endometrial WOI (rsERT) by three-time points sampling from the same patient at 48-hour intervals during one menstrual cycle. However, it is imperative to build a modified rsERT by single time point sampling in order to prevent multiple sampling and collateral harm.

**Methods:** A two-phase study was conducted. In the first phase, patients with successful clinical pregnancy after personalized embryo transfer (pET) guided by three-time points rsERT were recruited. Endometrial tissues obtained from single time point were used for the modified rsERT establishment. In the second phase, recurrent implantation failure (RIF) patients were recruited and assigned to experimental group underwent pET guided by modified rsERT’ or control group underwent conventional ET. Pregnant outcomes were recorded and analyzed.

**Results:** The modified rsERT was established using 91 eligible participants and could provide hour-based predictive result of endometrial WOI with an average accuracy of 94.51% with sensitivity and specificity being 92.73% and 96.27% using 10-fold CV. 176 RIF patients were recruited in the second phase (experimental group: n=88; control group: n=88). 40 of 88 (45.45%) patients showed WOI displacement, and 5.00% (2/40) of them were with advanced WOI, and the remaining 95.00% (38/40) with delayed WOI. The β-hCG positive rate, intrauterine pregnancy rate (IPR) and implantation rate (IR) of the experimental group were significantly improved (β-hCG positive rate: 67.05% vs. 39.77%, P=0.000; IPR: 61.36% vs. 31.82%, P=0.000; IR: 42.86% vs. 24.66%, P=0.001). While, pregnancy outcomes were not significantly different using different endometrial preparation protocols (β-hCG positive rate: 42.86% vs. 35.90%, P=0.508; IPR: 38.78% vs. 23.08%, P=0.116; IR: 30.12% vs. 17.46%, P=0.085).

**Conclusions:** The modified rsERT allowed WOI prediction using a single time point endometrial biopsy and pregnancy outcomes were significantly improved. This could provide an enhanced endometrial receptivity test as an alternative, requiring only a single time point sampling for RIF patients.

**Funding:** Research grants from Hunan Provincial Natural Science Foundation General Program (2023JJ30823) and Postdoctoral Fellowship Program of CPSF (GZC20233157).

## Introduction

A potentially developed embryo, endometrium in the receptive state and the synchronized molecular dialogue between them are three key processes for the successful pregnancy establishment (Teh, McBain, and Rogers 2016). In assisted reproductive technology (ART) scenario, about one-third of implantation failures can be ascribed to embryonic factor (Achache and Revel 2006). Maternal factors and endometrial-embryonic dysynchrony should be also considered when embryos fail to implant (Garneau and Young 2021). Even though there is heterogeneity in the definition of recurrent implantation failure (RIF), the asynchrony between embryo and endometrium may give rise to RIF (Sebastian-Leon et al. 2018).

After ovulation, the human endometrium transits from proliferation to secretory morphology and culminates with a spatially and temporally restricted period called the “window of implantation” (WOI). This self-limited period usually coincides with the 20th to 24th day of a regular menstrual cycle (Achache and Revel 2006), when the uterine milieu is favorable to blastocyst acceptance and implantation. However, the optimal duration of WOI is restrictedly short and only continues about 24-48 hours in human (Galliano et al. 2015). Profound evidences demonstrated that blastocyst-endometrial asynchrony would result in defective implantation or implantation failure (Ming et al. 2012; Healy et al. 2017; Omidi et al. 2021). Unsatisfactory pregnancy outcomes among RIF patients might be ascribed to the displacement of the WOI, in which embryos were transferred to the non-receptive endometrium at conventional timing (Hashimoto et al. 2017). Hence, accurate identification and prediction of WOI are essential to maximize the effectiveness of ART among infertile women who experienced RIF.

Currently, there are no reliable techniques to determine whether the endometrium is in synchrony with the embryo at the time of embryo transfer (ET). Since the advent of high-throughput “omics” techniques, series of genomic and transcriptomic diagnostic tools aiming to identify WOI have gained in popularity, including endometrial receptivity array (ERA) (Diaz-Gimeno et al. 2011), ER Map/ER Grade (Enciso et al. 2018) and the Win-Test (Haouzi et al. 2021). Previously, we have developed a novel RNA-sequencing-based endometrial receptivity test (rsERT) containing 175 genetic biomarkers to predict the WOI accurately (He et al. 2021). The three-time points sampling method from the same subject at 48-hour intervals during the same menstrual cycle (LH+5/LH+7/LH+9) allows a more precise identification of the optimal WOI. Subsequently, personalized embryo transfer (pET) guided by rsERT significantly improved the pregnancy outcomes of patients with RIF, especially those with WOI displacement (He et al. 2021). However, the three-time points sampling method can be costly and time-consuming, and more importantly, this invasive method renders patients more sensitive to pain.

To avoid multiple sampling and build an efficient WOI predictive tool, we first recruited participants to construct and optimize the rsERT and successfully established an estimation method by single time point sampling. Subsequently, a prospective controlled trial was conducted to validate its clinical benefit among RIF patients. This current study aims to provide a simpler and less invasive alternative method to identify the optimal time for embryo transfer among RIF patients.

## Materials and methods

### Study design and participants

This study was conducted at the Reproductive Medicine Center at the Xiangya Hospital of Central South University (CSU) in Changsha, Hunan, People’s Republic of China. All participants were undergoing ART between September 2018 and December 2021. This study involved two separate phases.

In the first phase (September 2018-June 2020), participants were recruited to optimize our previous predictive model for endometrial WOI, i.e., rsERT (He et al. 2021). Specifically, the inclusion criteria for allocated populations were as follows: 20-39 years of age; body mass index (BMI) 18-25 kg/m^2^; more than one cryopreserved transplantable cleavage-stage embryo or blastocyst. Endometrial tissues from three-time points sampling on days 5, 7 and 9 after the LH surge (LH+5, LH+7 and LH+9) or days 3, 5 and 7 after progesterone supplementation (P+3, P+5 and P+7) respectively were applied for endometrial WOI determination of the participants by our previous rsERT method (He et al. 2021). Patients with intrauterine pregnancy were recruited for the prototype of single time point model construction.

Afterwards, we used this prototype model to predict the receptive time of additional participants by single time point sampling at LH+7 or P+5. Those who successfully obtained intrauterine pregnancy after pET were further included to optimize the modified rsERT model. Here, intrauterine pregnancy was defined as the presence of a gestational sac with or without fetal heart activity in uterine cavity as evaluated by ultrasound 4-5 weeks after ET. Flowchart of phase 1 study was depicted in Figure 1.

**Figure 1.**
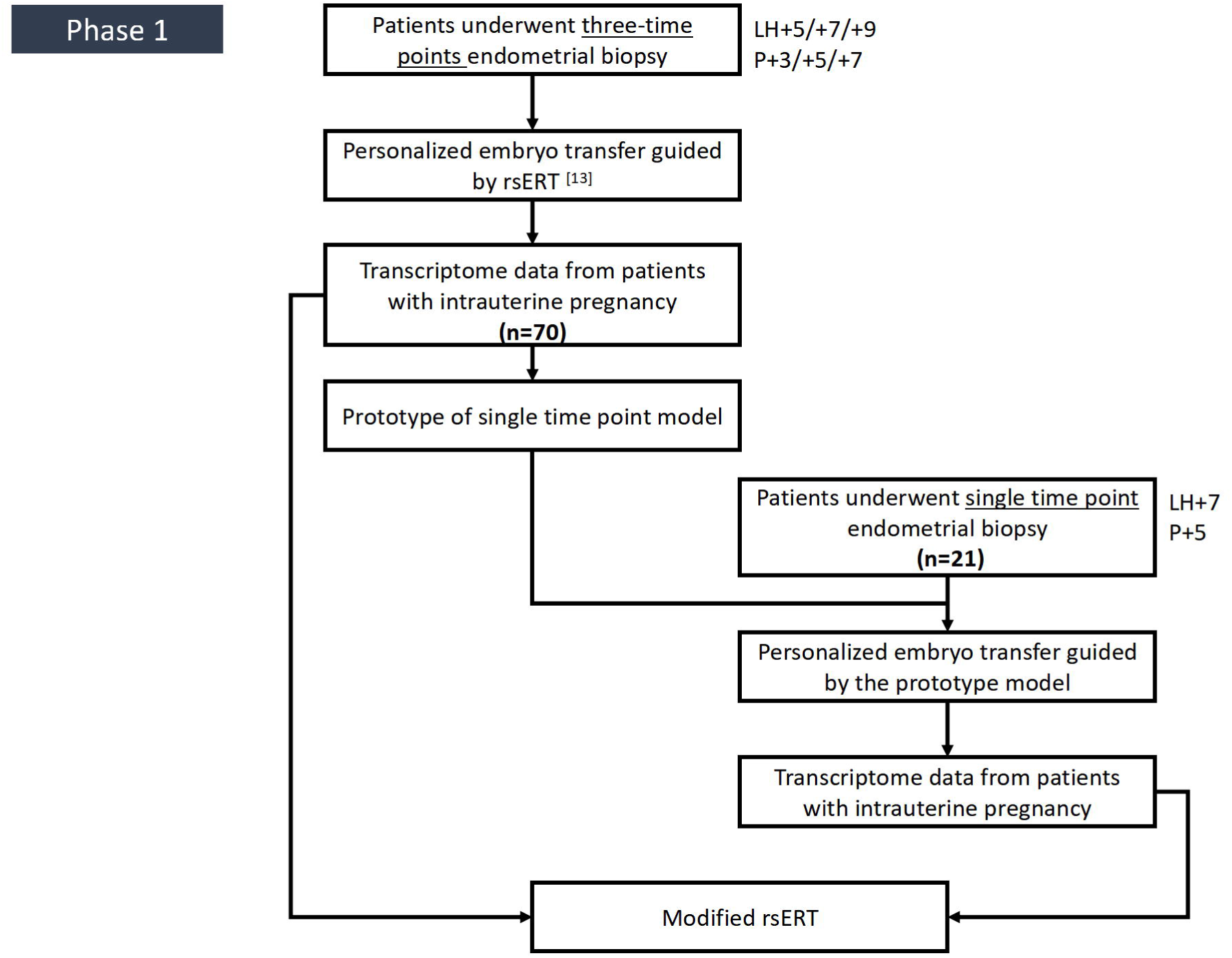
The flowchart of phase 1 study. Endometrial tissues from three-time points sampling on days 5, 7 and 9 after the LH surge (LH+5, LH+7 and LH+9) or days 3, 5 and 7 after progesterone supplementation (P+3, P+5 and P+7) respectively were applied for endometrial WOI determination of the participants by our previous rsERT method. Then, personalized embryo transfer guided by rsERT was conducted for each participant and 70 patients with intrauterine pregnancy were recruited for the prototype of single time point model construction. Then, 21 additional participants by single time point sampling at LH+7 or P+5 we used by this prototype model to predict the receptive time and those who successfully obtained intrauterine pregnancy after pET were further included to optimize the modified rsERT model.

In the second phase (June 2020-December 2021), participants were recruited to validate the clinical efficiency of modified rsERT among patients with RIF. This study was designed as a prospective controlled trial. Before recruitment, the sample size calculation in this part was performed under the condition of PASS software (version 15.0.5). Based on the results of the previous study (He et al. 2021), we assumed that the intrauterine pregnancy rate (IPR) was 60% in the experimental group and 30% in the control group. A two-sided P value is deemed statistically significant at P<0.05 and a power of 90%. Therefore, 53 subjects were required in each group. Considering a 10% loss-to-follow-up rate, 60 subjects in each group would be appropriate.

The inclusion criteria were as follows: 20-39 years of age; BMI 18-25kg/m^2^; and a history of RIF. RIF was defined as failure to achieve an intrauterine pregnancy after two consecutive cycles of fresh or frozen ET, in which the cumulative number of morphologically high-quality transferred embryos was no less than four for cleavage-stage embryos or two for blastocysts (Polanski et al. 2014). The criteria for high-quality embryos were as follows: cleavage-stage embryos: ≥7 blastomeres and <20% fragmentation on day 3 after fertilization (Alpha Scientists in Reproductive and Embryology 2011); blastocysts: ≥3BB on day 5 and day 6, graded based on the Gardner system (Gardner et al. 2019).

The exclusion criteria were as follows: endometrial abnormalities (e.g., intrauterine adhesions, endometrial polyps, endometrial hyperplasia, untreated chronic endometritis, and a thin endometrium); severe hydrosalpinx and did not receive tubal ligation or salpingectomy; submucous myoma, intramural hysteromyoma, or adenomyoma protruding towards the uterine cavity; endometriosis (stages III-IV); genital tuberculosis; uterine malformations; and other severe comorbidities (e.g., hypertension, diabetes mellitus, or malignant tumor).

After providing informed consent, patients who chose to receive the modified rsERT and subsequently performed pET were assigned to the experimental group. Those who decided not to receive rsERT and only underwent conventional ET belonged to the control group. Details of the phase 2 study were depicted in Figure 2.

**Figure 2.**
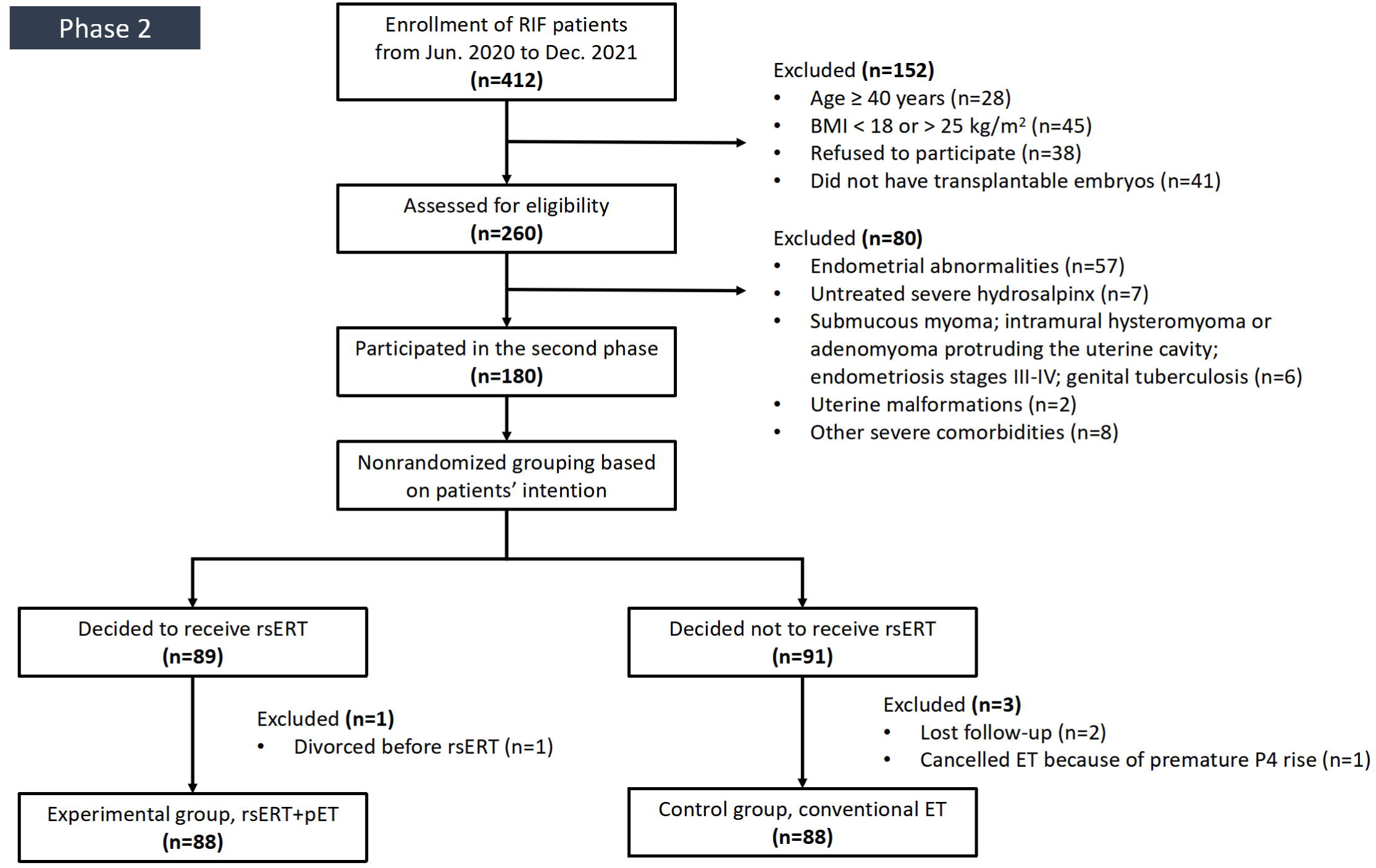
The flowchart of phase 2 study. 412 patients with RIF were initially recruited during June 2020-December 2021. Following careful inclusion criteria, 180 RIF patients were eligible to participate in the second phase to validate the clinical efficiency of modified rsERT. Then, the 180 patients were nonrandomized grouping into subgroup based on patient’s intention: the experimental group with 88 patients underwent modified rsERT and personalized embryo transfer (pET) and the control group with 88 patients underwent conventional embryo transfer (ET).

### Ethical approval

The study was approved by the Reproductive Medicine Ethics Committee of Xiangya Hospital, Central South University (CSU) (No. 2017002), and was registered in the Chinese Clinical Trial Registry (No. ChiCTR-DDD-17013375). Written informed consent was obtained from all participants and we conducted it according to the tenets of the Declaration of Helsinki.

### Endometrial biopsy, sample collection and processing

Written informed consent was obtained before sample collection. Endometrial tissues were collected using an endometrial sampler (AiMu Medical Science & Technology Co.; Liaoning, China). Timing of three-time points sampling method in natural cycle was on day 5, 7 and 9 after the LH surge (LH+5, LH+7 and LH+9) respectively. For hormone replacement therapy (HRT) cycle, estradiol administration was started on the third day of the menstrual cycle. Progesterone supplementation was started after at least 12 days of estrogen usage if the endometrium was >7mm and the endogenous P serum level was close to zero. The day of starting progesterone supplementation was considered P+0, and endometrial tissue was collected on day 3, 5 and 7 after progesterone supplementation (i.e., P+3, P+5 and P+7) respectively. The endometrium was only collected on LH+7 (nature cycle) or on P+5 (HRT cycle) if patient underwent single time point sampling.

Before sampling, the cervix was cleansed with saline. The tip of the endometrial sampler was placed into the uterine fundus, and 5-10mm^3^ of endometrial tissues were aspirated into the sampler. The collected endometrial tissues were immediately placed into 1.5ml RNAlater buffer (AM7020; Thermo Fisher Scientific, Waltham, MA, USA) for RNA stabilization, sealed, and cryopreserved at -20℃. Sequencing analysis was carried out within 7 days after sampling.

### RNA extraction, library construction and sequencing

The RNA-sequencing of endometrial biopsy samples was performed following the protocol described in previous study (He et al. 2021). In brief, total RNA extraction, quality control of RNA, reverse transcription and amplification, and (next-generation sequencing) NGS library construction were carried out by using commercial reagent kits. Then, under relevant parameters, single-end sequencing was performed on the HiSeq 2500 platform (Illumina, San Diego, CA, USA). The read length was set to 140 bp. The volume of raw data was approximately 5M reads.

### Candidate marker genes selection and predictive tool construction

Differentially expressed genes (DEGs) in different endometrial receptive conditions were screened according to the method in previous rsERT modeling (He et al. 2021). The expression values of these DEGs were then inputted as features for the random forest machine learning method to train the regression pattern of endometrial WOI time from each sample. Boruta R package (version 7.0.0) was applied to measure the importance of each feature (gene expression) (Kursa 2010). The features (genes) with confirmed importance were used for further model training and construction.

For optimal WOI point prediction, we applied the samples from three-time points sampling and single time point samples as training datasets for model construction and optimization. With the timing of endometrial biopsy, the corresponding predictive receptive results and subsequent clinical pregnancy outcomes, numerical value with hour precision of presumably optimal WOI in these training samples was defined. For example, if three samples from LH+5, LH+7 and LH+9 in one patient were predicted as pre-receptive, post-receptive and post-receptive by previous rsERT, blastocyst would be transferred on LH+6 (or day 3 cleavage embryo on LH+4). If patient obtained an intrauterine pregnancy afterwards, in this case, the numerical hour value for these three samples were roughly appointed as 24h, -24h and -72h, respectively.

Different combinations lead to 0h, 24h, -24h, 48h, -48h, 72h, -72h, 96h and -96h with 24-hour intervals. The random forest regression model from ranger R package (version 0.12.1) was therefore used to predict the optimal implantation timing with hour precision (Wright 2017). Infinitesimal jackknife resampling method was used to estimate the standard errors based on out-of-bag predictions of random forest strategy. The R-square value of model fitting and the 10-fold cross-validation approach was used to select model and evaluate the predictive performance of the model.

### pET guided by the modified rsERT and outcome measures

In the experimental group, pET was performed at the optimal WOI hour time predicted by the modified rsERT, corresponding to the timing of blastocysts transfer. Day 3 cleavage-stage embryos were transferred two days earlier accordingly. In the control group, conventional ET was performed as usually routine protocol without endometrial sampling. In this case, frozen-thawed cleavage-stage embryos were transferred on LH+5 or P+3, and blastocysts were transferred on LH+7 or P+5.

Patients were followed up to assess pregnancy outcomes as described below. Plasma β-human chorionic gonadotropin (β-hCG) was measured 12 days after ET. The intrauterine pregnancy and number of gestational sacs were assessed by ultrasound visualization 28 days after ET among patients with β-hCG positivity. The primary outcome measure was the IPR. A secondary outcome was the implantation rate (IR). IPR refers to the number of patients with intrauterine pregnancy per ET cycle. IR refers to the number of gestational sacs observed divided by the number of embryos transferred. Biochemical pregnancy was defined as a very early spontaneous abortion after a positive pregnancy had been determined and with a fall in plasma β-hCG concentration before the ultrasonic detection of gestational sacs (Bhatt et al. 2021).

### Statistical analysis

Statistical analysis was performed using IBM SPSS software (version 25.0, IBM Corp.). Continuous variables were described as the mean ± standard deviation (SD), or median and interquartile range (IQR) according to the distribution. Categorical variables were presented as frequency and percentage. The between-group differences among variables were analyzed by Student’s t-test or Mann-Whitney test, and Pearson’s chi-squared test or Fisher’s exact test for continuous and categorical variables, respectively. Values of P<0.05 for two-sided tests were considered statistically significant.

## Results

### Participants

In the first phase, from September 2018 to June 2020, endometrial biopsies from 91 eligible participants (mean±SD, age 31.32±3.59 years; BMI 20.93±1.96kg/m^2^) were finally recruited to construct and optimize the modified rsERT. The baseline characteristics of these 91 patients were shown in Table 1.

**Table 1.**
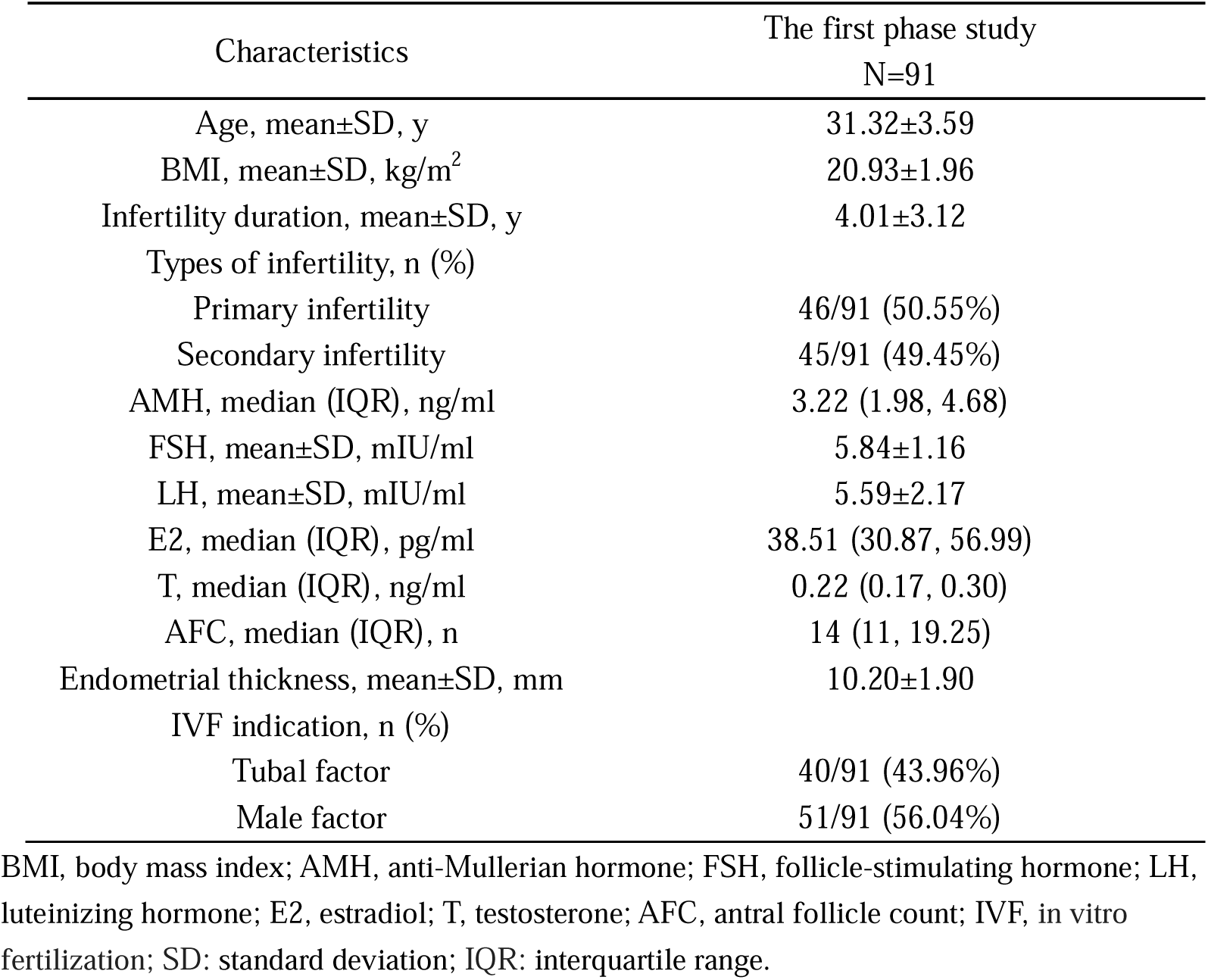
Demographic clinical characteristics of the participants for the modified rsERT establishment.

| Characteristics | The first phase study<br>N=91 |
| --- | --- |
| Age, mean±SD, y | 31.32±3.59 |
| BMI, mean±SD, kg/m<sup>2</sup> | 20.93±1.96 |
| Infertility duration, mean±SD, y | 4.01±3.12 |
| Types of infertility, n (%) |  |
| Primary infertility | 46/91 (50.55%) |
| Secondary infertility | 45/91 (49.45%) |
| AMH, median (IQR), ng/ml | 3.22 (1.98, 4.68) |
| FSH, mean±SD, mIU/ml | 5.84±1.16 |
| LH, mean±SD, mIU/ml | 5.59±2.17 |
| E2, median (IQR), pg/ml | 38.51 (30.87, 56.99) |
| T, median (IQR), ng/ml | 0.22 (0.17, 0.30) |
| AFC, median (IQR), n | 14 (11, 19.25) |
| Endometrial thickness, mean±SD, mm | 10.20±1.90 |
| IVF indication, n (%) |  |
| Tubal factor | 40/91 (43.96%) |
| Male factor | 51/91 (56.04%) |

BMI, body mass index; AMH, anti-Mullerian hormone; FSH, follicle-stimulating hormone; LH, luteinizing hormone; E2, estradiol; T, testosterone; AFC, antral follicle count; IVF, in vitro fertilization; SD: standard deviation; IQR: interquartile range.

In the second phase, considering the outpatient visit rate for RIF in our clinic, 176 eligible patients were finally enrolled during the clinical practice: 88 patients were included in the experimental group and 88 in the control group (Figure 2). The sample size in each group was more than that estimated by the PASS software (i.e., 60 vs. 60). Detailed clinical baseline characteristics were portrayed in Table 2. Baseline parameters, including age, BMI, infertility duration, types of infertility, number of previous failed cycles, main aetiology of infertility, the proportion of pre-implantation genetic screening/diagnosis (PGS/PGD) were all comparable in the experimental and control group (P>0.05). All patients were scheduled for cryopreserved embryo transfer in natural cycle or HRT cycle. There were no significant differences in the types of transferred cycle, endometrial thickness and endometrial types (P>0.05).

**Table 2.** Baseline characteristics of RIF patients in the experimental and control group.

| Characteristics | Experimental group<br>N=88 | Control group<br>N=88 | P value |
| --- | --- | --- | --- |
| Age (y) | 31.17±3.54 | 32.01±3.42 | 0.111 |
| BMI (kg/m<sup>2</sup>) | 20.71±1.82 | 21.25±2.24 | 0.078 |
| Infertility duration (y) | 4.69±3.12 | 4.85±3.45 | 0.749 |
| Types of infertility n (%) |  |  | 0.216 |
| Primary infertility | 58/88 (65.91%) | 50/88 (56.82%) |  |
| Secondary infertility | 30/88 (34.09%) | 38/88 (43.18%) |  |
| No. of previous failed cycles (n) | 2.92±1.10 | 2.75±1.02 | 0.287 |
| Main aetiology of infertility n (%) |  |  | 0.158 |
| Tubal | 48/88 (54.55%) | 64/88 (72.73%) |  |
| Ovulation disorder | 7/88 (7.95%) | 4/88 (4.55%) |  |
| Endometriosis | 6/88 (6.82%) | 3/88 (3.41%) |  |
| Diminished ovarian reserve | 7/88 (7.95%) | 8/88 (9.09%) |  |
| Male | 15/88 (17.05%) | 7/88 (7.95%) |  |
| Unexplained | 2/88 (2.27%) | 0/88 (0.00%) |  |
| Recurrent spontaneous abortion | 0/88 (0.00%) | 0/88 (0.00%) |  |
| Chromosomal or genetic abnormalities | 3/88 (3.41%) | 2/88 (2.27%) |  |
| PGS or PGD n (%) | 7/88 (7.95%) | 3/88 (3.41%) | 0.329 |
| Types of transferred cycle n (%) |  |  | 0.880 |
| Natural cycle | 40/88 (45.45%) | 39/88 (44.32%) |  |
| HRT cycle | 48/88 (54.55%) | 49/88 (55.68%) |  |
| Endometrial thickness (mm) | 9.58±1.92 | 9.89±1.67 | 0.265 |
| Endometrial types n (%) |  |  | 0.286 |
| A | 21/88 (23.86%) | 28/88 (31.81%) |  |
| B | 64/88 (72.73%) | 54/88 (61.36%) |  |
| C | 3/88 (3.41%) | 6/88 (6.83%) |  |

BMI, body mass index; PGS, pre-implantation genetic screening; PGD, pre-implantation genetic diagnosis; HRT, hormone replacement therapy.

### Model optimization and validation

The cut-off q-value of 0.001 was used to determine the DEGs among different endometrial statuses. After feature selection by random forest model, 201 biomarkers were finally selected for further model training and construction. Using the top 20 genes of high correlation between gene expression and different endometrial receptive statuses, we observed gradual (not sharp) genetic expression changes in the secretory phase of menstrual cycle (Figure 3). Therefore, the random forest regression model was fitted to predict optimal WOI time with hour precision. In other words, the modified rsERT could provide hour-based results. For instance, the 14 hours later, which indicates the timing of 14 hours later than endometrial biopsy time might be appropriate for ET to obtain a relatively higher possibility of embryo implantation. By adopting infinitesimal jackknife resampling method, the standard error and 95% confidence interval (CI) of predicted WOI time was obtained by the model.

**Figure 3.**
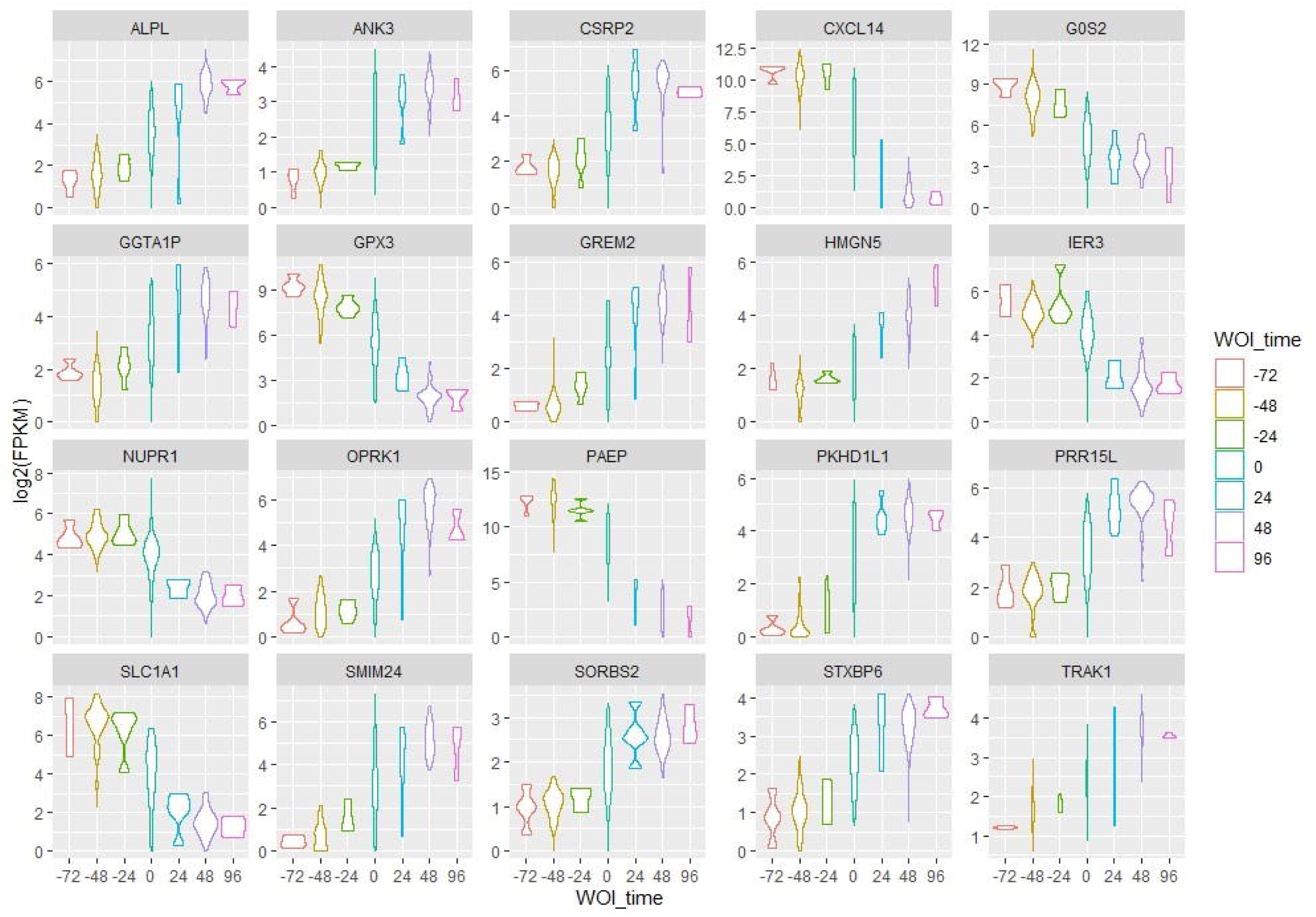
Gene expression pattern of the top 20 DEGs among different endometrial statuses. Gradual genetic expression changes were observed in the secretory phase of menstrual cycle.

Accordingly, the method is available to provide the window or range of optimal implantation time and guide clinical practice. 10-fold cross-validation was then applied to assess the performance of modified rsERT. A mean R-squared value of 0.92 was achieved, showing that 92% of the data fit the regression model. We found that the medians of the predicted mean WOI time in pre-receptive, receptive and post-receptive samples are approximately 48 hours, 0 hours and -48 hours respectively (Figure 4). Deviation points in Figure 4 are mostly from samples with WOI displacement. These data supported the notion that the modified rsERT had good performance in WOI prediction. With the cut-off values of -24 and 24 hours, we defined samples with predicted WOI time larger than 24 hours and less than -24 hours as pre-receptive and post-receptive status, receptively.

**Figure 4.**
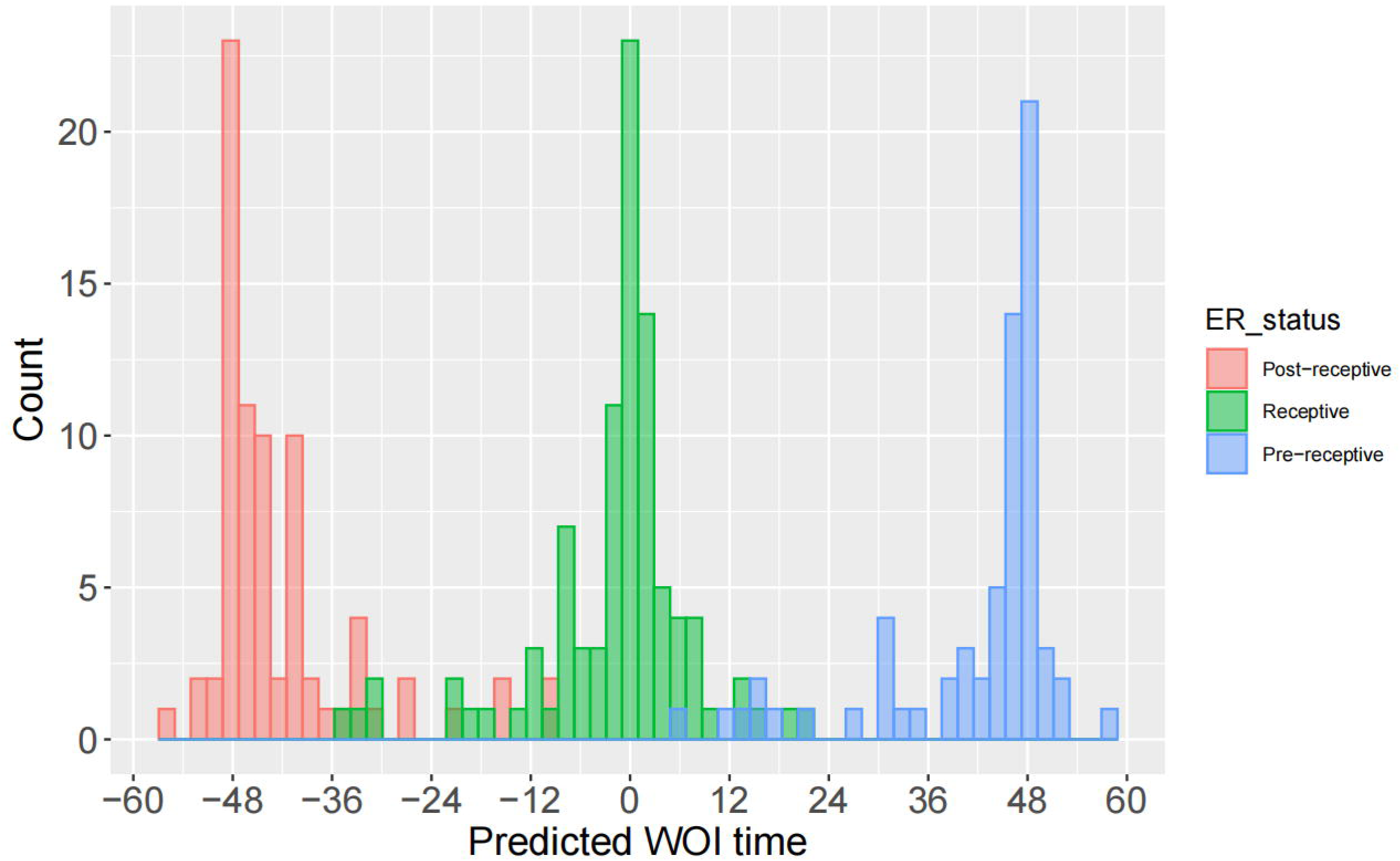
Correlation between predicted WOI time distribution and the corresponding endometrial receptive statuses. Medians of the predicted mean WOI time in pre-receptive, receptive and post-receptive samples are approximately 48 hours, 0 hour and -48 hours respectively. Deviation points are mostly from samples with WOI displacement.

Receptive samples were those predicted mean WOI time between -24 and 24 hours. Based on such standard, we obtained a mean 94.51% accuracy of this newly optimized rsERT model with sensitivity and specificity being 92.73% and 96.29%, respectively (Table 3). The current method allowed optimal WOI prediction by endometrial biopsy at single timing, thus avoiding second biopsy in the next menstrual cycle if the endometrial sample did not represent a receptive status.

**Table 3.** The performance of modified rsERT model with classification standard.

| Receptive status | Accuracy | Sensitivity | Specificity |
| --- | --- | --- | --- |
| Pre-receptivity | 94.53% | 89.06% | 100.00% |
| Receptivity | 93.56% | 95.70% | 91.43% |
| Post-receptivity | 95.44% | 93.42% | 97.45% |

Therefore, compared to the conventional ET, a more patient-friendly and cost-efficient individualized ET could be achieved by single time point sampling.

### Characteristics of WOI indicated by the modified rsERT and transferred embryos in the experimental and control group

During the second phase, a total of 88 NGS were constructed for RNA-sequencing by using endometrial biopsy samples from RIF patients in the experimental group, with a qualification rate of 100% (88 of 88). The results indicated WOI displacement in 40 of 88 (45.45%) (Table 4). Among them, advanced WOI occurred in two patients (2/40, 5.00%), and delayed WOI occurred in 38 patients (38/40, 95.00%).

**Table 4.** Characteristics of WOI indicated by modified rsERT and pregnancy outcomes in the experimental and control group.

| Characteristics | Experimental group<br>N=88 | Control group<br>N=88 | P value |
| --- | --- | --- | --- |
| WOI n (%) |  |  | / |
| Advanced | 2/88 (2.27%) | / |  |
| Delayed | 38/88 (43.18%) | / |  |
| On-time | 48/88 (54.55%) | / |  |
| Transferred embryo stage n (%) |  |  | 0.599 |
| D3 cleavage-stage embryos | 55/147 (37.41%) | 59/146 (40.41%) |  |
| D5 or D6 blastocysts | 92/147 (62.59%) | 87/146 (59.59%) |  |
| Good-quality embryo transferred rate<br>n (%) | 78/147 (53.06%) | 76/146 (52.05%) | 0.863 |
| No. of transferred embryo n | 1.67±0.47 | 1.66±0.48 | 0.874 |
| β-hCG positive rate n (%) | 59/88 (67.05%) | 35/88 (39.77%) | 0.000 |
| Biochemical pregnancy rate n (%) | 5/88 (5.68%) | 6/88 (6.82%) | 0.755 |
| IPR n (%) | 54/88 (61.36%) | 28/88 (31.82%) | 0.000 |
| IR n (%) | 63/147 (42.86%) | 36/146 (24.66%) | 0.001 |
| Ectopic pregnancy rate n (%) | 0/88 (0.00%) | 1/88 (1.14%) | 1.000 |

WOI, window of implantation; β-hCG, β-human chorionic gonadotropin; IPR, intrauterine pregnancy rate; IR, implantation rate.

There were no significant differences with respect to developmental stage of the transferred embryos (for D3: 37.41% vs. 40.41%; for D5 or D6: 62.59% vs. 59.59%, P=0.599), good-quality embryo transferred rate (53.06% vs. 52.05%, P=0.863) and number of transferred embryos (1.67±0.47 vs. 1.66±0.48, P=0.874) between the two groups (Table 4).

### Clinical efficiency of the modified rsERT-guided pET on pregnancy outcomes

Participants from the experimental group underwent pET according to the modified rsERT results and those from the control group conducted conventional ET respectively (Table 4). The β-hCG positive rate, IPR and IR of the experimental group were significantly higher than those in the control group (for β-hCG positive rate: 67.05% vs. 39.77%, P=0.000; for IPR: 61.36% vs. 31.82%, P=0.000; for IR: 42.86% vs. 24.66%, P=0.001). Biochemical pregnancy rate and ectopic pregnancy rate were comparable between the two groups (for biochemical pregnancy rate: 5.68% vs. 6.82%, P=0.755; for ectopic pregnancy rate: 0.00% vs.1.14%, P=1.000) (Table 4).

### Effects of endometrial preparation protocols on pregnancy outcomes of pET guided by the modified rsERT

To determine whether the natural cycle is superior to the HRT cycle, we performed a subgroup analysis of the experimental group. There were 40 populations in the natural cycle group and 48 in the HRT group. Baseline characteristics and pregnancy outcomes of these women are shown in Table 5. Age, BMI, infertility duration, types of infertility, number of previous failed cycles, main aetiology of infertility, the proportion of PGS or PGD, endometrial thickness, endometrial types, transferred embryo stage, good-quality embryo transferred and number of transferred embryos were comparable between the two groups (P>0.05). Moreover, there was no statistically significant difference regarding pregnancy outcomes between the two groups (P>0.05).

**Table 5.** Baseline characteristics and pregnancy outcomes of natural and HRT cycle in the experiment group.

| Characteristics | Natural cycle<br>N=40 | HRT cycle<br>N=48 | P value |
| --- | --- | --- | --- |
| Age (y) | 31.90±3.51 | 30.56±3.49 | 0.078 |
| BMI (kg/m<sup>2</sup>) | 20.49±1.89 | 20.89±1.75 | 0.311 |
| Infertility duration (y) | 4.65±3.45 | 4.73±2.86 | 0.908 |
| Types of infertility n (%) |  |  | 0.870 |
| Primary infertility | 26/40 (65.00%) | 32/48 (66.67%) |  |
| Secondary infertility | 14/40 (35.00%) | 16/48 (33.33%) |  |
| No. of previous failed cycles (n) | 3.08±1.23 | 2.79±0.97 | 0.240 |
| Main aetiology of infertility n (%) |  |  | 0.504 |
| Tubal | 23/40 (57.50%) | 25/48 (52.08%) |  |
| Ovulation disorder | 2/40 (5.00%) | 5/48 (10.42%) |  |
| Endometriosis | 2/40 (5.00%) | 4/48 (8.33%) |  |
| Diminished ovarian reserve | 2/40 (5.00%) | 5/48 (10.42%) |  |
| Male | 7/40 (17.50%) | 8/48 (16.67%) |  |
| Unexplained | 1/40 (2.50%) | 1/48 (2.08%) |  |
| Recurrent spontaneous abortion | 0/40 (0.00%) | 0/48 (0.00%) |  |
| Chromosomal or genetic abnormalities | 3/40 (7.50%) | 0/48 (0.00%) |  |
| PGS or PGD n (%) | 5/40 (12.50%) | 2/48 (4.17%) | 0.238 |
| Endometrial thickness (mm) | 10.01±2.20 | 9.23±1.59 | 0.065 |
| Endometrial types n (%) |  |  | 0.778 |
| A | 11/40 (27.50%) | 10/48 (20.83%) |  |
| B | 28/40 (70.00%) | 36/48 (75.00%) |  |
| C | 1/40 (2.50%) | 2/48 (4.17%) |  |
| Transferred embryo stage n (%) |  |  | 0.069 |
| D3 cleavage-stage embryos | 30/66 (45.45%) | 25/81 (30.86%) |  |
| D5 or D6 blastocysts | 36/66 (54.55%) | 56/81 (69.14%) |  |
| Good-quality embryo transferred rate n (%) | 39/66 (59.09%) | 39/81 (48.15%) | 0.186 |
| No. of transferred embryo n | 1.65±0.48 | 1.69±0.47 | 0.102 |
| β-hCG positive rate n (%) | 26/40 (65.00%) | 33/48 (68.75%) | 0.709 |
| Biochemical pregnancy rate n (%) | 2/40 (5.00%) | 3/48 (6.25%) | 1.000 |
| IPR n (%) | 24/40 (60.00%) | 30/48 (62.50%) | 0.810 |
| IR n (%) | 28/66 (42.42%) | 35/81 (43.21%) | 0.924 |

BMI, body mass index; PGS, pre-implantation genetic screening; PGD, pre-implantation genetic diagnosis; β-hCG, β-human chorionic gonadotropin; IPR, intrauterine pregnancy rate; IR, implantation rate.

Genetic expression patterns were also explored between samples obtained from the natural and HRT cycles. With the same cut-off q-value as 0.001, no DEGs were observed (Figure 5), implying little influences of endometrial preparation protocols on the efficiency of this modified model.

**Figure 5.**
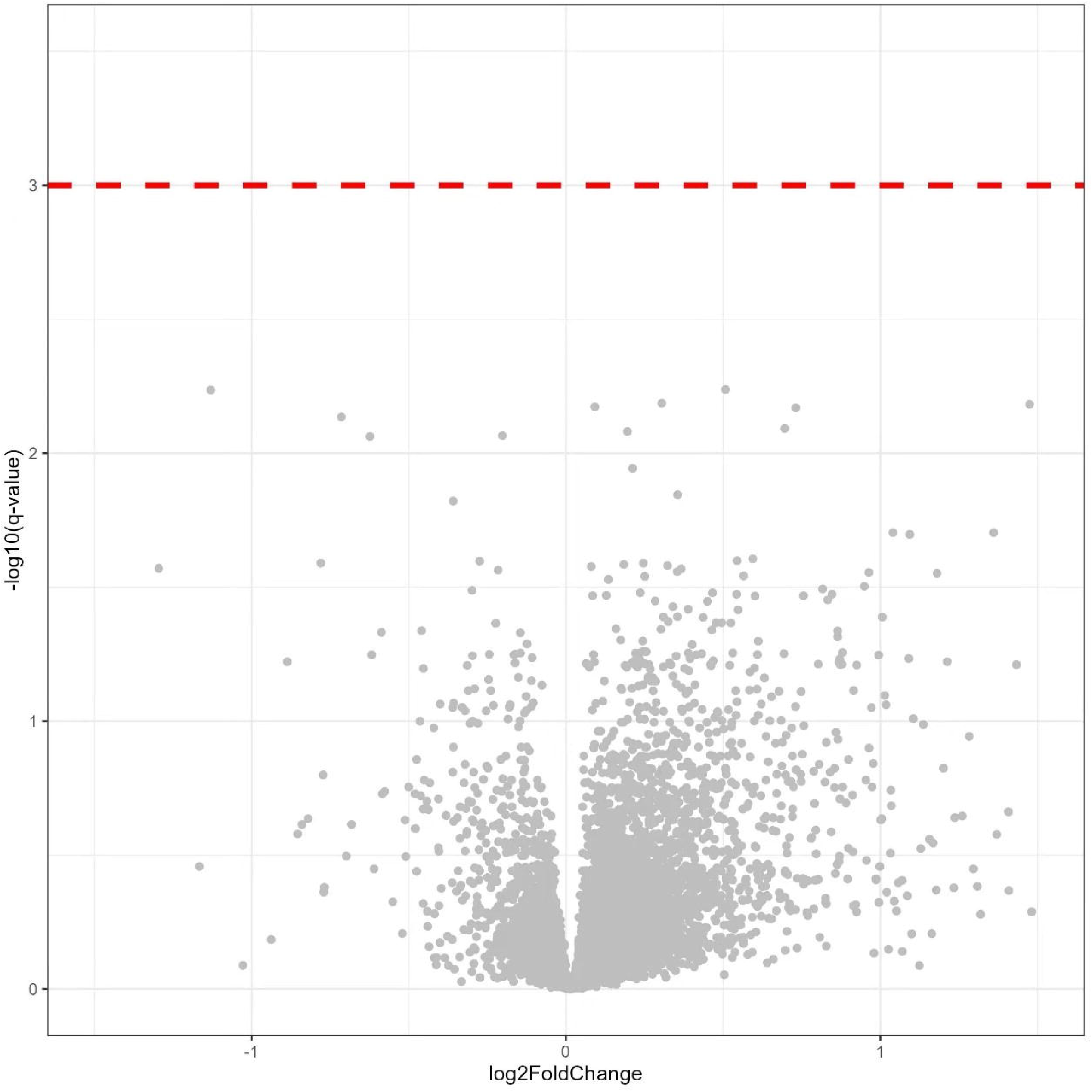
Volcano plot of differential transcriptomic profile between samples obtained from the natural cycle and HRT cycle. No differentially expressed genes were observed between samples obtained from natural and HRT cycle.

### Effects of endometrial preparation protocols on pregnancy outcomes of conventional ET

We further evaluated the effects of different endometrial preparation regimens on pregnancy outcomes after conventional ET. There were 39 participants who underwent FET in the natural cycle and 49 in the HRT cycle. Baseline characteristics including age, BMI, infertility duration, types of infertility, number of previous failed cycles, main aetiology of infertility, and others were comparable between the two groups (P>0.05, Table 6). With respect to individuals who performed conventional ET without endometrial sampling, the β-hCG positive rate, IPR and IR in the HRT group were slightly higher than those in the natural cycle group, whereas differences were not statistically significant (for β-hCG positive rate: 42.86% vs. 35.90%, P=0.508; for IPR: 38.78% vs. 23.08%, P=0.116; for IR: 30.12% vs. 17.46%, P=0.085; Table 6).

**Table 6.**
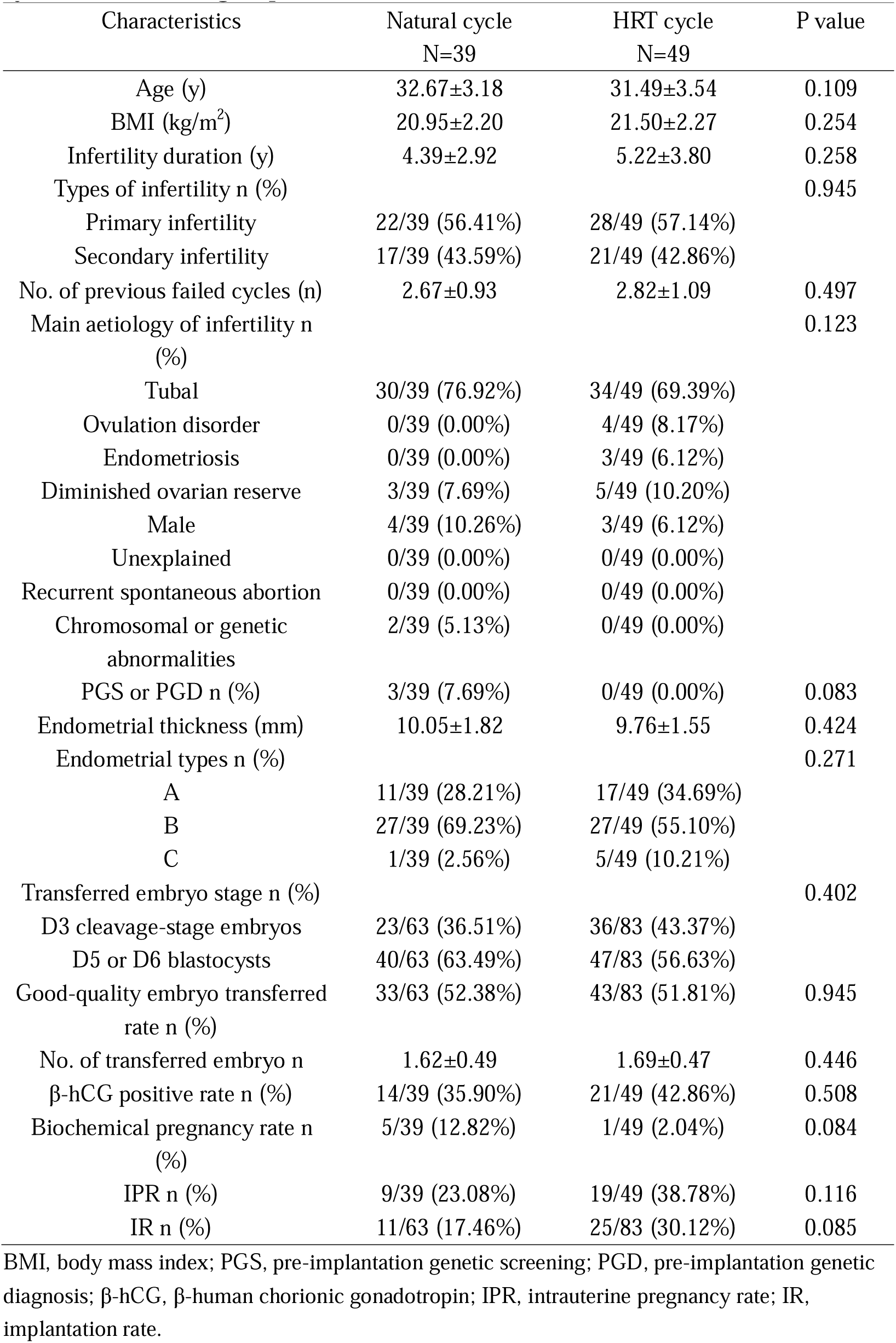
Baseline characteristics and pregnancy outcomes of natural and HRT cycle in the control group.

| Characteristics | Natural cycle<br>N=39 | HRT cycle<br>N=49 | P value |
| --- | --- | --- | --- |
| Age (y) | 32.67±3.18 | 31.49±3.54 | 0.109 |
| BMI (kg/m<sup>2</sup>) | 20.95±2.20 | 21.50±2.27 | 0.254 |
| Infertility duration (y) | 4.39±2.92 | 5.22±3.80 | 0.258 |
| Types of infertility n (%) |  |  | 0.945 |
| Primary infertility | 22/39 (56.41%) | 28/49 (57.14%) |  |
| Secondary infertility | 17/39 (43.59%) | 21/49 (42.86%) |  |
| No. of previous failed cycles (n) | 2.67±0.93 | 2.82±1.09 | 0.497 |
| Main aetiology of infertility n (%) |  |  | 0.123 |
| Tubal | 30/39 (76.92%) | 34/49 (69.39%) |  |
| Ovulation disorder | 0/39 (0.00%) | 4/49 (8.17%) |  |
| Endometriosis | 0/39 (0.00%) | 3/49 (6.12%) |  |
| Diminished ovarian reserve | 3/39 (7.69%) | 5/49 (10.20%) |  |
| Male | 4/39 (10.26%) | 3/49 (6.12%) |  |
| Unexplained | 0/39 (0.00%) | 0/49 (0.00%) |  |
| Recurrent spontaneous abortion | 0/39 (0.00%) | 0/49 (0.00%) |  |
| Chromosomal or genetic abnormalities | 2/39 (5.13%) | 0/49 (0.00%) |  |
| PGS or PGD n (%) | 3/39 (7.69%) | 0/49 (0.00%) | 0.083 |
| Endometrial thickness (mm) | 10.05±1.82 | 9.76±1.55 | 0.424 |
| Endometrial types n (%) |  |  | 0.271 |
| A | 11/39 (28.21%) | 17/49 (34.69%) |  |
| B | 27/39 (69.23%) | 27/49 (55.10%) |  |
| C | 1/39 (2.56%) | 5/49 (10.21%) |  |
| Transferred embryo stage n (%) |  |  | 0.402 |
| D3 cleavage-stage embryos | 23/63 (36.51%) | 36/83 (43.37%) |  |
| D5 or D6 blastocysts | 40/63 (63.49%) | 47/83 (56.63%) |  |
| Good-quality embryo transferred rate n (%) | 33/63 (52.38%) | 43/83 (51.81%) | 0.945 |
| No. of transferred embryo n | 1.62±0.49 | 1.69±0.47 | 0.446 |
| β-hCG positive rate n (%) | 14/39 (35.90%) | 21/49 (42.86%) | 0.508 |
| Biochemical pregnancy rate n (%) | 5/39 (12.82%) | 1/49 (2.04%) | 0.084 |
| IPR n (%) | 9/39 (23.08%) | 19/49 (38.78%) | 0.116 |
| IR n (%) | 11/63 (17.46%) | 25/83 (30.12%) | 0.085 |

BMI, body mass index; PGS, pre-implantation genetic screening; PGD, pre-implantation genetic diagnosis; β-hCG, β-human chorionic gonadotropin; IPR, intrauterine pregnancy rate; IR, implantation rate.

## Discussion

For nearly 20 years, transcriptomic analysis on ER has been developed drastically (Messaoudi et al. 2019). Several diagnostic tools for endometrial dating have been commercialized to personalize the FET, e.g., ERA (Diaz-Gimeno et al. 2011), ER Map/ER Grade (Enciso et al. 2018) and the Win-Test (Haouzi et al. 2021). Previously, we established an RNA-sequencing-based endometrial receptivity test (rsERT) via three-time points sampling and demonstrated its clinical effectiveness afterwards (He et al. 2021). However, either multiple time points sampling of previous rsERT or potential second sampling risk in another menstrual cycle of other WOI predictive models may impede their clinical applications. In order to identify endometrial WOI more simply and accurately, this current study modified the rsERT which recently allows optimal WOI estimation via single time point endometrial sampling.

Based on classifications of rsERT from three-time points biopsies and their corresponding clinical outcomes, we constructed an endometrial WOI diagnostic tool (i.e., the modified rsERT) for single time point sampling. The three-time points’ sequencing data in the same cycle showed varied endometrial transcriptomic profiles from pre-receptive to the post-receptive phase, providing a fundamental basis for available predictive results which were accurate to the hour. By adopting RNA-sequencing and random forest algorithm, 201 biomarker genes were selected for the modified rsERT model. Interestingly, the variation trend of these genes during the sampling period was mainly continuous. Only a tiny number of genes showed abrupt change. According to the single-cell transcriptomic study, human endometrial WOI opens with an abrupt and discontinuous transcriptomic activation, and closes with continuous transitions (Wang et al. 2020). The discrepancies might be due to the varied interval of endometrial biopsy and interference from other types of cells in our tissue samples.

Herein, we listed top 20 genes among the 201 biomarkers as they revealed significant differences in expression among different endometrial receptivity statuses. Those genes are mainly involved in immune response and inflammatory process (PAEP, CXCL14, OPRK1 and NUPR1)(Pathare, Zaveri, and Hinduja 2017; Yu et al. 2020), anti-oxidative stress (GPX3), cellular proliferation, apoptosis and differentiation (G0S2, CSRP2)(Welch et al. 2009; Kihara et al. 2011), osteogenesis (ALPL, GREM2)(Wang et al. 2017), cytoskeletal anchor activity (ANK3)(Wang et al. 2016) and glutamate transportation (SLC1A1). Half of the top 20 genes, namely ALPL, ANK3, CSRP2, CXCL14, GPX3, GREM2, IER3, SLC1A1, OPRK1 and PEAP, overlap with the 238 biomarkers identified by the study of ERA (Diaz-Gimeno et al. 2011).This result showed that there was biologically significant consistency of biomarkers identified in both ERA and the modified rsERT.

It is acknowledged that there are differences between ERA and rsERT with respect to the study design, sample collection and detection methods as we previously described (He et al. 2021). For example, samples were obtained from the same patient at 48-hour intervals within the same menstrual cycle to build the rsERT model, in contrast to the ERA where samples were obtained from different women on different cycle days (Diaz-Gimeno et al. 2011). Besides, we defined the receptive period as day LH+7 or P+5 combining with a subsequent intrauterine pregnancy, which is more reliable than the previous determination with LH+7 or P+5 alone. In this case, the DEGs and biomarkers might vary from different studies, and their biological functions have not been fully explained yet. Much effort will put into the functional investigation of these genes by using cutting-edge technologies (e.g., single-cell RNA sequencing) in the near future.

Aiming to demonstrate its clinical efficiency, the modified rsERT-guided pET was performed among RIF patients. The proportion of a displaced WOI identified by the modified rsERT was 45.45% (40/88), similar to the result determined by ERA reported elsewhere (Saxtorph et al. 2020), indicating a higher WOI displacement rate in this specific populations. Moreover, the modified rsERT model also enables predictive statement accurate to hours for blastocyst transfer, e.g., the recommended optimal time for ET is 36 (95% CI 30-42) hours after sampling. As such, β-hCG positivity rate, IPR, and IR were significantly higher in the experimental group compared with the control group. Although some studies challenged the clinical utility of pET among patients with a history of failed transfers or RIF (Cozzolino et al. 2020; Eisman et al. 2021), significantly improved pregnancy rate and implantation rate were observed when the timing of ET was properly adjusted according to the displaced WOI (Ruiz-Alonso et al. 2013; He et al. 2021; Haouzi et al. 2021). Some aspects including experimental design, inclusion and exclusion criteria of the recruitment and the testing platform may explain the inconsistencies.

In addition, it is worth noting that differential expression analysis of biomarker genes reveals no significant difference between the endometrial samples from natural and HRT cycles (Figure 5). This suggests that the feature selection using a machine learning algorithm could eliminate the discrepancy between these two groups. Altmäe et al. demonstrated that when compared to the HRT-FET cycle among women with RIF, the whole endometrial gene expression profile of RIF patients at the WOI in natural cycle was more similar to the pattern of the fertile controls (Altmae et al. 2016). One possible reason for yielding a contradictory result is that the cutoff value in Altmäe’s study is 0.05, whereas ours is stricter, i.e., 0.001. Besides, Altmäe et al. did not report the clinical results of the two groups. Interestingly, in the present study, pregnancy outcomes did not differ significantly between natural cycle and HRT cycle among RIF patients. Therefore, this predictive model would be appropriate for patients undergoing both natural and HRT cycles. Meanwhile, we noticed a slightly higher rates of β-hCG positivity, clinical pregnancy and embryo implantation among participants underwent an HRT-FET protocol in the control group, and the between-group difference was not significant partly due to the relatively small sample size.

Even though the mechanism of embryo implantation during WOI remains a “black box” (Franasiak et al. 2016), rsERT provides an optimal timing for pET strategy. In fact, except for WOI displacement, the duration of WOI is a critical but easily overlooked factor. For people with narrower WOI, the endometrium allows a shorter time window for embryo implantation, in which case the timing of ET seems more crucial. For patients with an extremely narrow WOI, the modified rsERT could provide an optimal timing (to the hour) for ET, in case embryos were transferred beyond the duration of their WOI. In other words, rsERT offered individualized ET according to their personal WOI, whether it was displaced or not.

Nevertheless, caution is warranted due to the non-randomized design and limited sample size. Even though baseline characteristics and other parameters were comparable between the two cohorts, a more valid conclusion should be drawn from the randomized controlled trial (RCT) accomplishment. Our preliminary clinical results from this prospective cohort study would lay the foundation of future RCT. Besides, embryonic factors should be taken into consideration as well. In fact, we have already designed a multi-center RCT of the modified rsERT in combination with PGT to validate its clinical application value further.

## Conclusions

To sum up, we modified our previous predictive model for endometrial WOI (rsERT), allowed WOI prediction via single time point endometrial biopsy. When compared to the RIF participants underwent conventional ET, pregnancy outcomes had been improved among patients after pET guided by the modified rsERT. The results indicate that we could provide patients with an improved effective endometrial receptivity test that only requires single time point sampling.

## Data Availability

The datasets used and/or analysed during the current study are available from the corresponding author upon reasonable request.

## Abbreviations

ART: assisted reproductive technology
AMH: anti-Mullerian hormone
AFC: antral follicle count
BMI: body mass index
CI: confidence interval
DEGs: differentially expressed genes
ER: endometrial receptivity
ERA: endometrial receptivity array
ET: embryo transfer
E2: estradiol
FSH: follicle-stimulating hormone
FET: frozen embryo transfer
HRT: hormone replacement therapy
IPR: intrauterine pregnancy rate
IR: implantation rate
IQR: interquartile range
IVF: in vitro fertilization
LH: luteinizing hormone
P: progesterone
pET: personalized embryo transfer
PSM: propensity score matching
PGS: pre-implantation genetic screening
PGD: pre-implantation genetic diagnosis
RIF: recurrent implantation failure
rsERT: RNA-sequencing based endometrial receptivity test
SD: standard deviation
T: testosterone
WOI: window of implantation
β-hCG: β-human chorionic gonadotropin.

## Acknowledgements

The authors are extremely grateful to all the volunteers for participation in this study.

## Authors’ contributions

QZ and YL designed research and carefully revised the manuscript; AH and TY wrote the paper; SL, YZ and CW participated in the model optimization; JZ, NL, DL, YL, YW, BX, JH and SX executed the trial. JF, HL and HW collected and analyzed the data. All authors read and approved the final manuscript.

## Declarations

### Consent for publication

Not applicable.

